# Effects of food insecurity on infant feeding confidence and practices during the COVID-19 pandemic and its waves

**DOI:** 10.1101/2025.02.07.25321881

**Authors:** Maha Hussain, Jessica Hyun, Elena Arduin, Margaret H. Kyle, Wanda Abreu, Tessa Scripps, Adrita Khan, Melissa Glassman, Melissa S. Stockwell, Lauren Walzer, Dani Dumitriu, Cristina R. Fernández

## Abstract

**Background:** The COVID-19 pandemic has intensified economic hardships, with potential negative impacts on food insecurity and infant feeding beliefs and practices. The relationship between food insecurity and infant feeding beliefs and practices during the pandemic is not yet fully understood. Neither is how these relationships changed over the course of the waves of the pandemic. We examined these relationships in a cohort of infants born during the various waves of the COVID-19 pandemic in New York City (NYC).

**Methods:** We conducted a cross-sectional analysis of infants enrolled from birth into the COVID-19 Mother-Baby Outcomes (COMBO) study and born March 2020 to May 2024. We measured food insecurity in the prior 30 days with a 2-item survey adapted from Hunger Vital Sign™, infant feeding confidence at hospital discharge, and current infant feeding practices.

**Results:** In our sample, 40% of women had been exposed to prenatal SARS-CoV-2 infection and approximately 24% of mothers were food insecure. There was a significant association between food insecurity and prenatal SARS-CoV-2 infection, Spanish as one’s preferred language, and self-identifying as Latina. In unadjusted models of the entire sample, food insecurity was associated with formula feeding, with 44% of food insecure mothers opting to formula feed vs. 28% of food secure mothers (p<0.001), but this relationship was no longer significant after adjusting for covariates (p=0.059). In comparing the first and second waves of the pandemic (March 2020-December 2021 vs. January 2022-May 2024), there was no significant difference in rate of food insecurity. When comparing different waves of the pandemic, food insecurity was associated with increased likelihood of formula feeding, even after adjusting for confounders.

**Discussion:** Food insecurity was initially associated with feeding methods, but this relationship lost significance after adjusting for confounders. However, when analyzed separately, food insecurity was significantly linked to lower odds of exclusive breastfeeding during different waves of the pandemic, suggesting the influence of external factors like policy changes and social support variations. Other factors, such as maternal BMI, ethnicity, and delivery mode, were also significantly associated with breastfeeding practices, highlighting the need for targeted interventions to support breastfeeding, especially among food-insecure mothers.

## Introduction

Food insecurity is a global public health challenge that was exacerbated by the SARS-CoV-2 pandemic.^1^ Food insecurity is defined as when nutritionally adequate and safe foods are unavailable or when the ability to acquire such foods in socially acceptable ways is limited or uncertain.^2^ Though food insecurity has always been a challenge in the United States, rates of household food insecurity in the nation have increased dramatically from 11% in 2019 to 38% in March 2020.^3,4^ In April 2020, 35% of households with children under 18 reported experiencing food insecurity.^4^ During this period, record-high unemployment rates, supply shortages, income insecurity, and related factors not only had independent impacts but also interacted in complex ways, likely contributing to the significant rise in food insecurity observed during the pandemic, with many households experiencing food insecurity for the first time^5,6^ Food insecurity is associated with household stressors that may impact feeding of the most vulnerable members of the household, namely children and infants. The effects of food insecurity and feeding this vulnerable group, especially in the context of a global pandemic, are not yet fully understood.

Breastfeeding continues to be regarded as an optimal form of infant feeding.^7,8^ Experts from the World Health Organization recommend exclusive breastfeeding for infants up to 4-6 months of age, followed by continued breastfeeding alongside appropriate complementary foods until the age of 2 years.^9^ Breastfeeding confers significant health benefits, while decreased breastfeeding rates are often associated with poorer maternal and infant health outcomes.^10,11^ Yet only 25.8%^12^ of women in the United States meet recommendations to exclusively breastfeed for the first 6 months of a child’s life.^13,14^ Several studies have documented changes in breastfeeding rates during the pandemic. For example, a study completed in the United Kingdom found that 18.9% of mothers reported they stopped breastfeeding due to lack of support, which was more common amongst mothers from minority backgrounds.^15^ Another study compared breastfeeding patterns between pandemic and pre-pandemic groups and found that women in the pandemic group were 15% less likely to exclusively breastfeed.^16^

Using data from the COVID-19 Mother Baby Outcomes (COMBO) longitudinal study, we previously found that mothers with prenatal SARS-CoV-2 infection were less likely to breastfeed at 1, 2, and 4 months of infant age than mothers without prenatal SARS-CoV-2 infection.^17^ This difference was particularly evident in mothers who delivered during the second wave of the pandemic, and was present in our sample despite our hospital system’s encouragement of newborns rooming-in with their mothers and direct breastfeeding, contrary to early national health recommendations. Women of Latina ethnicity who had a SARS-CoV-2 infection during pregnancy were particularly likely to report decreased breastfeeding rates.^17^ These results highlight the need to protect and increase traditional networks of breastfeeding support that may have been disrupted during the COVID-19 pandemic, and to increase accessibility of these resources to women in sociodemographic groups historically underrepresented in healthcare.

Decreased breastfeeding rates have been associated with higher levels of household food insecurity,^18–20^ potentially contributing to perceived poorer maternal diet, high stress levels, and the limiting of healthy foods.^21^ Initial recommendations during the beginning of the COVID-19 pandemic included mother-newborn separation and no direct breastfeeding. However, due to lack of evidence for vertical transmission, providers began to encourage mother-infant skin-to-skin contact and direct breastfeeding after appropriate hand and breast hygiene.^22^ Though it appears that breastfeeding rates decreased during the acute phases of the pandemic, it is not yet well understood how these relationships developed as the social environment changed throughout its various waves.

Though the economic and social disruptions caused by the COVID-19 pandemic are well-documented, it is unclear how infant feeding practices may have been influenced by food insecurity during this time. As a result of the COVID-19 pandemic, parents experienced heightened levels of stress, with mothers likely bearing a disproportionate share of the burden, which may have affected infant feeding practices.^23–28^ Further understanding of the associations between food insecurity in the context of the COVID-19 pandemic and maternal-infant feeding practices is crucial to inform programs and interventions that seek to improve breastfeeding rates and support mother-infant bonding. This is especially important when considering historically marginalized populations that are disproportionally burdened by food insecurity and disparities in breastfeeding rates. As New York City (NYC) was an early epicenter of the pandemic and since a majority of our population resides in NYC’s low-income Washington Heights’ neighborhood primarily composed of Black and Latino-identifying individuals, it is essential to explore and understand these relationships.

The primary objectives of our study were to determine the association between household food insecurity and breastfeeding practices and maternal feeding confidence during the COVID-19 pandemic in NYC. Secondary objectives of our study were to examine this association and food insecurity and breastfeeding practices between the early and later time periods of the COVID-19 pandemic. We hypothesized that mothers with food insecurity during the pandemic would be less likely to breastfeed and report lower infant feeding confidence.

## Methods

### Study design, study site, and study population

Our cross-sectional study was nested within Columbia University Irving Medical Center’s (CUIMC) COVID-19 Mother Baby Outcomes (COMBO) ongoing prospective cohort study, investigating the health of mother-infant dyads with prenatal maternal severe acute respiratory syndrome coronavirus 2 (SARS-CoV-2) infections versus case-matched negative controls. History of maternal SARS-CoV-2 infection was determined by nasopharyngeal PCR beginning March 22, 2020 and additionally by serological testing for SARS-CoV-2 antibodies beginning July 20, 2020. Mother-infant dyads with history of maternal SARS-CoV-2 infection (exposed) were matched to 1-3 control dyads (unexposed) with a comparable infant sex, gestational age at birth, mode of delivery and date of birth who tested negative for SARS-CoV-2 at the time of delivery and had no record of maternal SARS-CoV-2 infection before or during pregnancy. Dyads enrolled into COMBO at infants’ birth to 2 months of age. Participants were recruited by research assistants and clinicians via phone calls, SMS, and emails, and then were asked to complete a 30-60-minute self-administered survey. Eligibility criteria for our study include COMBO-enrolled English-and/or Spanish-speaking mothers who gave birth from March 2020 at CUIMC/NewYork-Presbyterian Hospital (NYP).

CUIMC/NYP is located in the Washington Heights/Inwood community of northern Manhattan and handles 6,000-7,000 live births per year. Forty-six percent of residents of this community are foreign born, a rate higher than the national average of 13.7%, and 15.5% live in poverty.^29,30^

All study procedures were approved by the Columbia University Irving Medical Center Institutional Review Board.

### Exposure Variable

The main predictor variable—food insecurity in the prior 30 days—was measured using a 2-item survey, adapted from the validated Hunger Vital Sign™.^31^ The 2-item survey captured both acute worry of having enough food (“Within the past 1 month have you worried that your food would run out before you got money to buy more?”) and not having resources to acquire food (“Within the past 1 month did the food you bought just not last and you didn’t have money to get more?”). Participants answered either “Often,” “Sometimes,” or “Never” to both questions. Participants responded to this question upon COMBO baseline enrollment. Mothers who answered “often” or “sometimes” to either or both items were defined as being food insecure.

### Outcome Measures

Main outcomes of (i) maternal confidence with infant feeding after hospital discharge and (ii) infant feeding practices were measured via a self-administered baseline questionnaire at COMBO enrollment at either 1-or 2-months postpartum. These questions were developed by clinicians at CUIMC working in the Newborn Clinic at the onset of the pandemic for the purpose of data collection for the COMBO study. Breastfeeding practices were measured from an infant feeding practice question where mothers reported: “all breastmilk,” “mostly breastmilk but some formula,” “equal amounts of breastmilk and formula,” “mostly formula but some breastmilk,” or “all formula.”^32^ The 5-level variable was subsequently combined into a 3-level variable of “all breastmilk,” “breastmilk and formula” and “all formula.” Maternal confidence with infant feeding (both formula and breastfeeding) was measured using a 4-item Likert scale with responses ranging from “completely confident” to “not at all confident.” Maternal prenatal breastfeeding intention was also measured by an affirmative response to the questions: “Has it always been your plan to do mostly/all breastmilk?”

### Covariates

Additional sociodemographic covariates associated with food insecurity and breastfeeding practices *a priori* were abstracted from electronic health records: maternal age, infant age, self-reported ethnicity and race, insurance status, gestational age, delivery method, and infant sex based on results from other studies.^21,33–36^

### Statistical analysis

Mother-infant dyads with complete exposure and outcome data were included in the final analytic sample. Bivariate analyses compared covariates and outcomes by food insecurity status. Categorical variables were analyzed using Chi-square and Fisher Exact tests and continuous variables were analyzed using Mann-Whitney U tests. Ordinal logistic regression models estimated the association between food insecurity and outcome measures significant on bivariate analyses. After assessing for collinearity among variables, models were adjusted for non-collinear *a priori* covariates and covariates significant at the p < 0.25 level during model-building: maternal prenatal SARS-CoV-2 infection status, main language spoken in the home, infant sex, infant age at COMBO enrollment, and maternal age. Secondary analyses compared food insecurity level of maternal infant feeding confidence levels between waves of the NYC COVID-19 pandemic (Wave 1 March 2020-December 2021 and Wave 2 January 2022-May 2024). The adjusted models were then stratified by wave. Statistical analyses were conducted using RStudio (Version 2024.04.2+764, R Core Team, 2025), and statistical significance was at the P < 0.05 level.

## Results

Our sample was comprised of N=606 mother-infant dyads who gave birth between March and May of 2024, of which 40% of dyads had been exposed to prenatal SARS-CoV-2 infection. Sample descriptive statistics are summarized in Table 1. Half (48% [N=289]) of mothers self-identified as Latina, 33% as White (N=200), 10% as Black (N=63), and 4% as Asian (N=27).

**Table 1.** Cohort Characteristics.

| Table 1. Cohort Characteristics |  |  |  |  |
| --- | --- | --- | --- | --- |
| Variable<br>No. (%) or Mean (± Standard<br>Deviation) | Overall<br>Sample<br>(N=606) | Food<br>Insecure<br>(N=144) | Food<br>Secure<br>(N=446) | P-value |
| <u>Maternal Characteristics</u> |  |  |  |  |
| SARS-CoV-2 Exposure |  |  |  |  |
| Exposed | 244 (40%) | 60 (42%) | 161 (36%) | 0.041 |
| Unexposed | 289 (48%) | 55 (38%) | 233 (52%) |  |
| Unknown | 73 (12%) | 29 (20%) | 52 (12%) |  |
| Language |  |  |  |  |
| English | 464 (77%) | 77 (54%) | 377 (85%) | p<0.001 |
| Spanish | 109 (17%) | 64 (45%) | 39 (9%) |  |
| Other | 22 (4) | 1 (1%) | 21 (5%) |  |

| Ethnicity |  |  |  |  |
| --- | --- | --- | --- | --- |
| Latino or Hispanic or Spanish Origin | 289 (48%) | 118 (83%) | 171(38%) | p<0.001 |
| Not Latino or Hispanic or Spanish Origin | 290 (48%) | 24 (17%) | 266 (60%) |  |
| Race (Asian) |  |  |  |  |
| Asian | 27 (4%) | 1 (1%) | 25 (6%) | p<0.001 |
| Black or African American | 63 (10%) | 17 (12%) | 43 (10%) |  |
| White | 200 (33%) | 22 (15%) | 176 (40%) |  |
| Other | 305 (50%) | 102 (71%) | 193 (43%) |  |
| Medical Coverage |  |  |  |  |
| Public | 279 (46%) | 115 (80%) | 150 (34%) | p<0.001 |
| Private | 316 (52%) | 27 (19%) | 287 (64%) |  |
| Maternal Age at Delivery | 31.6 (± 5.7) | 30.0 (± 6.2) | 32.2 (± 5.5) | p<0.001 |
| Maternal BMI at Delivery | 31.8 (± 6.7) | 32.1 (± 5.5) | 31.8 (± 7.1) | 0.24 |
| Maternal Educational Level |  |  |  |  |
| 7-12<sup>th</sup> grade | 26 (4%) | 16 (11%) | 10 (2%) | p<0.001 |
| High school degree/GED | 42 (7%) | 15 (10%) | 27 (6%) |  |
| Partial college | 52 (9%) | 20 (14%) | 32 (7%) |  |
| 2-year college degree/4-year college degree/Trade school/apprenticeship | 154 (25%) | 36 (25%) | 117 (26%) |  |
| Graduate degree | 154 (25%) | 14 (10%) | 140 (31%) |  |
| Other | 4 (1%) | 1 (1%) | 3 (1%) |  |
| Infant Characteristics |  |  |  |  |
| Baby Sex |  |  |  |  |
| Male | 329 (54%) | 87 (60%) | 233 (52%) | 0.11 |
| Female | 277 (46%) | 57 (40%) | 213 (48%) |  |
| Mode of Delivery |  |  |  |  |
| Vaginal | 375 (62%) | 77 (53%) | 286 (64%) | 0.029 |
| C-section | 231 (36%) | 67 (47%) | 160 (36%) |  |
| Number of Children in Household |  |  |  |  |
| 1 | 163 (27%) | 27 (19%) | 135 (30%) | 0.008 |
| 2-3 | 156 (26%) | 43 (30%) | 113 (25%) |  |
| 4+ | 32 (5%) | 12 (8%) | 19 (4%) |  |
| Gestational Age | 38.7 (± 1.9) | 38.6 (± 1.7) | 38.7 (± 1.8) | 0.34 |
| **Birth Weight in Grams** | 3255.3 (± 557.6) | 3269.6 (±571.7) | 3258.4 (± 543.9) | 0.78 |

Exposed: *in utero* maternal SARS-CoV-2 exposed infants born during the pandemic; Unexposed: Unexposed infants born during the pandemic; Stress Controls: Infants born pre-pandemic in February 2020.
\*Comparisons performed using Chi Squared Test for binomial variables with expected cell values of five and over, Fisher's Exact Test for binomial variables with expected cell values under five, and Mann-Whitney U test for continuous variables.
\*Some data not available for all participants due to nonresponse.

Mean maternal age was 31.6 ± 5.7 years, mean maternal body mass index (BMI) at delivery was 31.8 ± 6.7 kg/m^2^, and 36% (N=231) mothers delivered via C-section. Mean gestational age for all infants was 38.7 ± 1.9 weeks. More than half of the mothers (52%, N=316) had private insurance.

Bivariate statistics comparing the relationship between food insecurity status and demographic characteristics are also summarized in Table 1. Approximately 24% of mothers were food insecure in the last 30 days based on the criteria of the validated Hunger Vital Sign criteria.^31^ There was a significant association between food insecurity and prenatal SARS-CoV-2 infection, with mothers who experienced prenatal SARS-CoV-2 infection more likely to experience food insecurity in the last month (p=0.04). Spanish-speaking mothers (45% food insecure mothers vs. 9% of food secure Spanish-speaking mothers, p<0.001) and mothers who self-identified as of Latina (83% vs. 17% of non-Latina food insecure mothers, p<0.001) were more likely to experience food insecurity. Mothers experiencing food insecurity were also more likely to be younger (mean age 30.0 ± 6.2 years vs. 32.2 ± 5.5 years, p<0.001), have public health coverage (80% of food insecure mothers had public health coverage, while 19% had private coverage, p<0.001), have lower levels of education (p<0.001), and have more children in their households (p=0.01). Mothers who delivered via C-section were also more likely to experience food insecurity (p=0.03).

Approximately 54% of the entire sample reported being completely confident in feeding their infant after hospital discharge. Food insecurity was not significantly associated with maternal infant feeding confidence (p=0.062). There was no significant difference between food insecure and food secure mothers’ intention to breastfeed. 70% of food insecure mothers reported an intention to breastfeed vs. 75% of food secure mothers.

In unadjusted models of the entire sample, food insecurity was associated with formula feeding (p<0.001). Only 18% of mothers experiencing food insecurity exclusively breastfed their child, while 26% exclusively formula-fed. By contrast, 41% of mothers not experiencing food insecurity exclusively breastfed their child, while 13% of them only formula-fed.

Although not statistically significant, after adjusting for covariates, mothers experiencing food insecurity had approximately twice the odds of formula feeding compared to food-secure mothers (adjusted odds ratio [AOR] of exclusively breastfeeding 0.47, 95% confidence interval - 1.54, 0.03, p=0.059) (Table 2).

**Table 2.** Infant Feeding Method, Adjusted OR and CI: Modelling the Adjusted Odds of Formula Feeding vs. Breastfeeding.

|  | OR | 95% CI | P-value |
| --- | --- | --- | --- |
| **<u>Reference Group</u>** |  |  |  |
| **Food Insecurity Status (Food Secure)** |  |  |  |
| Food Insecure | 0.47 | -1.54, 0.030 | 0.059 |
| **SARS-CoV-2 Infection (Unexposed)** |  |  |  |
| Exposed | 0.58 | -1.22, 0.13 | 0.12 |
| **Maternal Ethnicity (Not of Latino or Hispanic or Spanish Origin)** |  |  |  |
| Latino or Hispanic or Spanish Origin | 0.21 | -2.30, -0.85 | <0.001 |
| **Infant's Adjusted Age** | 1.43 | 0.020, 0.70 | 0.039 |
| **Maternal BMI** | 1.09 | 0.031, 0.14 | 0.0022 |
| **Mode of Delivery (Vaginal)** |  |  |  |
| C-section | 0.48 | -1.44, -0.051 | 0.035 |

**Table 3.** Infant Feeding Method, Adjusted OR and CI: Modelling the Adjusted Odds of Formula Feeding vs. Breastfeeding.

|  | OR | 95% CI | P-value |
| --- | --- | --- | --- |
| **<u>Reference Group</u>** |  |  |  |
| **Food Insecurity Status (Food Secure)** |  |  |  |
| Food Insecure | 0.266 | -1.98, -0.68 | <0.001 |
| **Wave** |  |  |  |
| Wave 1 | 12.0 | 0.75, 5.42 | 0.021 |

**Table 4.**
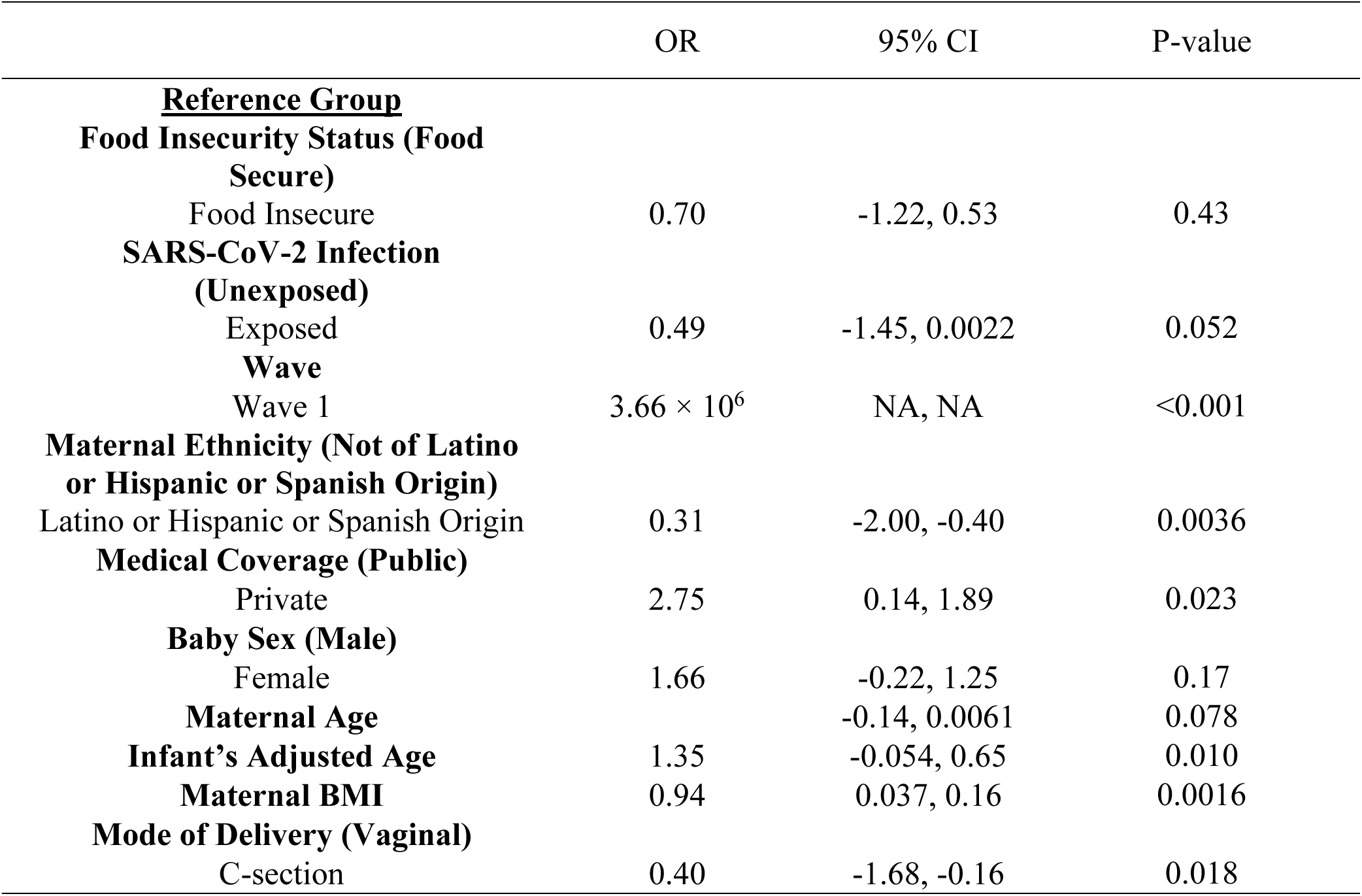
Infant Feeding Method, Adjusted OR and CI: Modelling the Adjusted Odds of Formula Feeding vs. Breastfeeding.

**Table 4.** Infant Feeding Method, Adjusted OR and CI: Modelling the Adjusted Odds of Formula Feeding vs. Breastfeeding by Wave.

|  | Wave 1 |  |  | Wave 2 |  |  |
| --- | --- | --- | --- | --- | --- | --- |
|  | OR | 95% CI | P-value | OR | 95% CI | P-value |
| **<u>Reference Group</u>** |  |  |  |  |  |  |
| **Food Insecurity Status (Food Secure)** |  |  |  |  |  |  |
| Food Insecure | 0.33 | -1.87, -0.34 | 0.0044 | 0.034 | -6.74, -1.14 | 0.012 |
| **Infant's Adjusted Age** | 1.37 | -0.018, 0.65 | 0.063 | 0.18 | -3.57, -0.25 | 0.036 |
| **Maternal BMI** | 1.11 | 0.047, 0.16 | <0.001 | 1.18 | 0.054, 0.32 | 0.0095 |
| **Maternal Age** | 0.89 | -0.18, -0.049 | <0.001 | 0.88 | -0.28, -0.0051 | 0.049 |
| **Mode of Delivery (Vaginal)** |  |  |  |  |  |  |
| C-section | 0.39 | -1.68, -0.25 | 0.0087 | 0.26 | -3.10, 0.24 | 0.11 |

In the adjusted model that includes food insecurity and COVID-19 wave, the interaction was significant (p=0.021). In comparing the first and second waves of the pandemic (March 2020-December 2021 vs. January 2022-May 2024), there was no significant difference in rate of food insecurity (p=0.09) or in mothers’ confidence in their ability to feed their infant after hospital discharge (p=0.74). Mothers who gave birth in the first wave of the pandemic were more likely to exclusively breastfeed compared to others who gave birth in the second wave of the pandemic (38% vs. 24%, p=0.004). Food insecurity was significantly associated with less of a likelihood of formula feeding in both waves, with ORs of 0.33 (95% CI:-1.87,-0.34, p=0.0044) in Wave 1 and 0.034 (95% CI:-6.74,-1.14, p=0.012) in Wave 2. This suggests that food-insecure mothers are more likely to formula feed.

Infant’s adjusted age shows a positive but non-significant association with formula feeding in Wave 1 (OR=1.37, p=0.063), whereas in Wave 2, it is significantly associated with lower odds of formula feeding (OR=0.18, p=0.036), indicating that as infants grow older, they may be more likely to continue breastfeeding. Maternal BMI is consistently associated with increased formula feeding across both waves, with significant ORs of 1.11 (p<0.001) in Wave 1 and 1.18 (p=0.0095) in Wave 2, highlighting that higher maternal BMI may be a predictor of formula feeding. Additionally, maternal age is inversely associated with formula feeding, with significant results in both waves (p<0.001 in Wave 1, p=0.049 in Wave 2), suggesting older mothers are more likely to breastfeed. Mode of delivery (C-section) is linked to a lower likelihood of formula feeding in Wave 1 (OR=0.39, p=0.0087), but the effect is not statistically significant in Wave 2 (p=0.11), indicating that the impact of delivery method on feeding choice may diminish over time.

## Discussion

Almost one-third of the mothers in our sample experienced food insecurity, a rate higher than the national household food insecurity rate of 11% from 2019,^4^ suggesting that the pandemic may have exacerbated rates of food insecurity in New York City, consistent with reports of increased rates of food insecurity across the nation. In 2022, food insecurity rates were reported to be 16.6% in the Washington Heights and Inwood area.^37^ Though most would associate breastfeeding as a way of saving money, reports from food insecure mothers in Canada found that breastfeeding did not actually save families money as there was often little money for food to begin with in these households, meaning that infant feeding method had little to no effect on finances.^38^ Food insecure mothers reported that breastfeeding was still costly for them, as they had to purchase groceries to ensure that their breastmilk provided their children with all of the necessary nutrients. Mothers also feared that their own inability to prioritize their nutrition would negatively affect their children. In Nova Scotia, the cost for the extra food needed for a lactating mother (between the ages 19 to 30) cost $35.33 per month on top of the cost for a basic nutritious diet, which was $222.47 in 2012.^38^ Increased financial insecurity during the pandemic^39^ may have exacerbated these concerns.

In the overall sample of mothers who delivered an infant during the first and second waves of the COVID-19 pandemic in New York City, food insecurity was significantly associated with feeding method, but was no longer significantly associated after adjusting for confounders. However, it is important to note that these results trended towards significance (p=0.059). Though food insecurity only appears to trend towards significance, Maternal Ethnicity, Infant’s Adjusted Age, Maternal BMI, and Mode of Delivery all were significantly associated with exclusive breastfeeding practices. By contrast, secondary analyses determined that food insecurity was significantly associated with lower adjusted odds of exclusive breastfeeding in Wave 1 and Wave 2 when the waves were analyzed separately. This suggests that possibility of external factors not captured by the study, such as changes in NYC and governmental policies, during Wave 1 and Wave 2 that may have dampened the relationship between food insecurity status and infant feeding across the entire time period of the overall sample.

Buccini et al. conducted a meta-analysis of 12 studies and found that household food insecurity was associated with reduced rates of exclusive breastfeeding.^40^ In this meta-analysis, the study with the largest sample size (N=7950),^33^ found that women with food security were more likely to exclusive breastfeed their child. Yet previous studies have found no association between food insecurity and exclusive breastfeeding duration and practices or with other poor feeding behaviors.^20,33,36,41^ During other stressful conditions, such as natural disasters and conflict situations, mothers reported that lacking privacy, spaces conducive to breastfeeding, limited fluid and energy intake, stress, and exhaustion deterred them from breastfeeding.^42,43^ Mothers may have similar concerns in the context of a pandemic. Work-from-home and stay-at-home orders may have conferred less privacy to mothers seeking to breastfeed. Moreover, increased access to certain resources such as infant formula through programs like WIC, which saw a 2.1% increase in use during the first year of the pandemic, and other food emergency programs may have disincentivized women from exclusive breastfeeding.^44,45^

The COVID-19 pandemic’s effects on breastfeeding practices are yet to be fully understood. Mothers in our sample with confirmed SARS-CoV-2 infection were less likely to breastfeed. Studies suggest that COVID-19 is not transmitted through breastmilk,^46^ yet multiple studies found that mothers with SARS-CoV-2 infection were less likely to breastfeed their child or have reduced breastfeeding duration.^47,48^ Some studies found overall lower breastfeeding rates during the pandemic,^46^ while others reported increased breastfeeding duration due to the pandemic.^49^ Decreased breastfeeding rates could be a result of increased stressors during the pandemic, while work-from-home opportunities and stay-at-home orders may have increased the amount of time parents spent near their children, thus providing opportunities for breastfeeding.

Women from our sample who identified as Latino or Hispanic or Spanish Origin were also less likely to exclusively breastfeed. Other studies have found that minority women were less likely to continue breast feeding during the pandemic, citing a lack of face-to-face support.^50^ Services like breastfeeding support can result in additional expenses that individuals more likely to experience socioeconomic stressors, like women of Latino or Hispanic or Spanish origin,^51^ may be unable to afford, which may have negatively impacted their ability to secure the supports needed to increase their breastfeeding rates and duration. Moreover, the COVID-19 pandemic drastically limited both the overall availability of services and in-person service opportunities, as well as options for families to connect in-person with extended relatives, all of which may have contributed to decreased breastfeeding rates.

Mothers with lower BMI were more likely to breastfeed. Previous studies have similarly seen that higher BMI levels can adversely impact breastfeeding initiation and duration.^52,53^ Women with higher BMIs often experience lactogenesis and an increase in medical conditions that may prevent them from breastfeeding.^53^ Women with higher BMIs are more likely to belong to social groups who are less likely to breastfeed (such as those of lower socio-economic status) and experience greater body image dissatisfaction, both of which have been associated with lower breastfeeding rates.^53^ Mothers who delivered via C-section were less likely to breastfeed, which has also been seen in other studies,^54,55^ due to problems with latching, positioning and more pain when compared to women who delivered vaginally.^55^ The maternal and fetal stress responses associated with complications during C-section delivery may be related to increased difficulties in breastfeeding, while lactogenesis may also be affected by the surgery.^55^

By contrast to our hypothesis, we did not find significant relationship between food insecurity and maternal infant feeding confidence. Maternal feeding confidence has been associated with perception of control during labor,^56^ past performance (e.g., past breastfeeding experiences with other children), vicarious experiences (e.g., watching other women breastfeed), verbal persuasion (e.g., encouragement from influential others, such as friends, family, and lactation consultants), and physiological responses (e.g., fatigue, stress, and anxiety).^57^ Food insecure mothers were more likely to have more children; this increased parity, which implies increased feeding experience, may have been a stronger driver of their infant feeding confidence than current experiences of economic hardship with food insecurity. People experiencing food insecurity are more likely to be unemployed^58^, meaning they may have had more time to spend time with family and friends, thereby potentially engaging in more vicarious experiences and verbal persuasion, and have increased opportunities for breastfeeding.

As previously discussed, several studies have found a relationship between food insecurity and lack of breastfeeding.^40^ Though food insecurity only appeared to trend towards significance in our overall sample, interestingly it was a significant factor when analyzing data in the context of the different waves of the pandemic. These results indicate that maternal factors, food security status, and delivery mode all play roles in feeding choices, with significant shifts between time points throughout the COVID-19 pandemic. The observed significant association between food insecurity and infant feeding practices in the wave-specific analyses, but not in the overall sample, suggests potential temporal variations in the relationship between economic hardship and breastfeeding behaviors. One possible explanation is that external factors, such as changes in social support, government assistance programs, or pandemic-related disruptions, may have influenced feeding decisions differently across time points. Additionally, the stratification of data into waves may have allowed for the detection of nuanced patterns that were obscured in the pooled analysis, where heterogeneity across waves could dilute statistical significance. It is also possible that different sample characteristics, such as maternal stress levels or access to lactation support, varied between waves and impacted breastfeeding continuation disproportionately. Similar to our overall sample, maternal BMI was consistently associated with increased odds of formula feeding in both waves, suggesting that maternal health and body composition may play a role in feeding decisions. Prior studies have indicated that higher maternal BMI is linked to physiological and psychological barriers to breastfeeding, including experiencing delayed lactogenesis, perceived insufficient milk supply, and lower breastfeeding self-efficacy.^59^ These findings highlight the multifaceted nature of infant feeding choices and the need to better understand the factors that influence them and the urgent need for targeted interventions to support breastfeeding.

Here we present food insecurity prevalence and key breastfeeding behaviors and infant feeding outcomes among postpartum mothers during the first waves of the COVID-19 pandemic in New York City, one of the earliest pandemic epicenters in the U.S. Limitations of our study include the cross-sectional design and the sample from a single hospital system, limiting generalizability to other health systems and other regions of the U.S. that experienced first waves of the COVID-19 pandemic later than NYC. Similarly, our sample size may limit the power to identify small effect sizes. The sample included self-reported baseline questionnaires from mothers with infants between 0-4 months-old, which had the potential to allow for self-reporting biases. Unlike the validated Hunger Vital Sign™ that measures food insecurity in the last 12 months, our adapted food insecurity measure only captured acute food insecurity experiences in the prior month, which does not account for food insecurity experiences from earlier time points.

Next steps include longitudinal assessment of food insecurity across multiple time points in the ongoing COMBO study, and collection of quantitative and qualitative data around prenatal, perinatal, and postpartum maternal feeding expectations and maternal-infant feeding experiences to support the development of interventions to support breastfeeding initiation, continuation, and maternal confidence. Further research into how already existing health disparities such as food insecurity and low breastfeeding effects are affected in the context of the pandemic will also be beneficial in understanding this complex topic. Future implications of our research include informing prenatal and postpartum interventions to increase breastfeeding knowledge, education, community support and resources and to increase breastfeeding among women with food insecurity and other environmental hardship experiences. Future research should also consider longitudinal approaches to disentangle the dynamic effects of food insecurity on breastfeeding and explore potential policy interventions that may mitigate these disparities.

## Data Availability

All data produced in the present study are available upon reasonable request to the authors.

---

**Supplemental Table 1.**
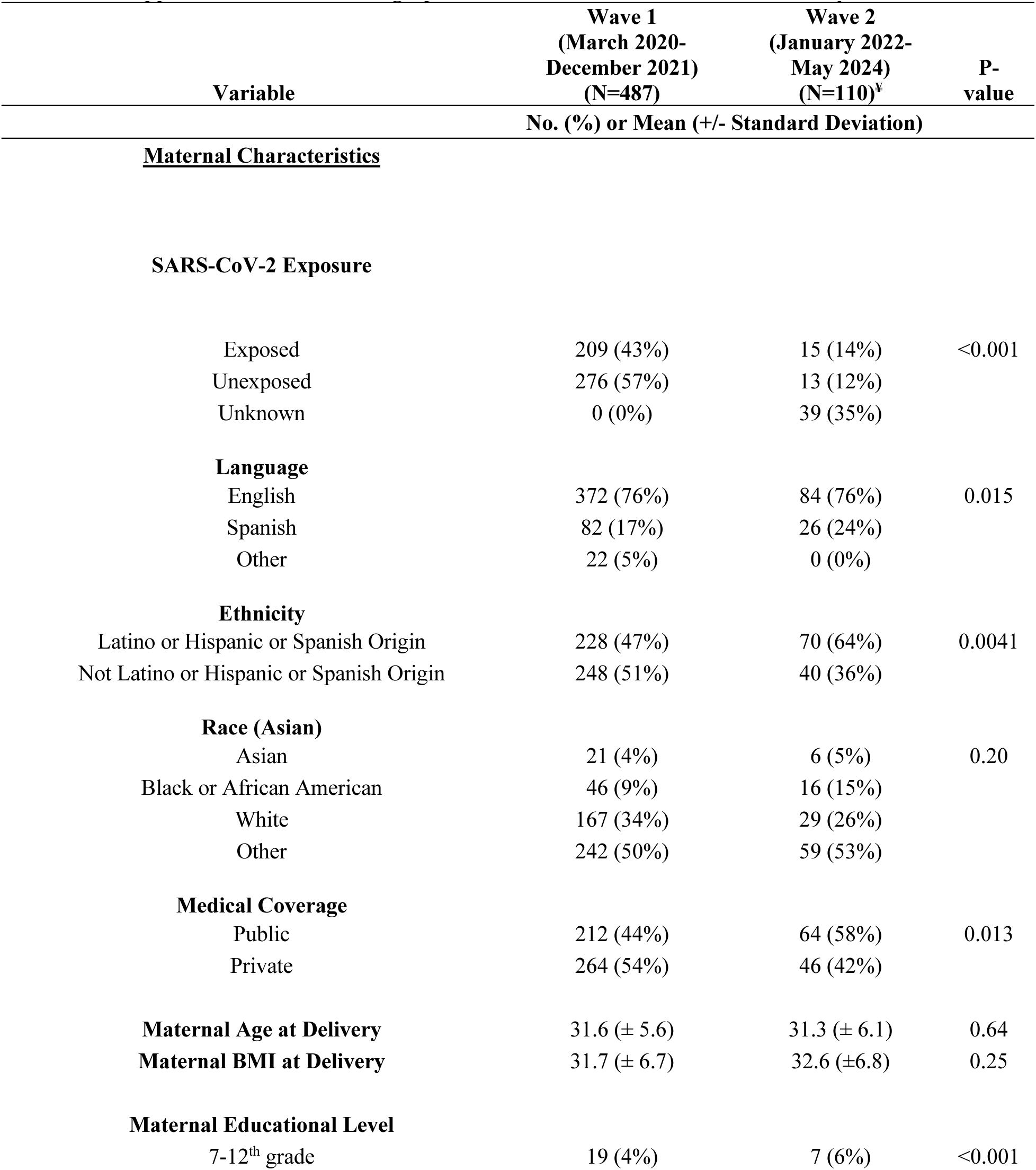
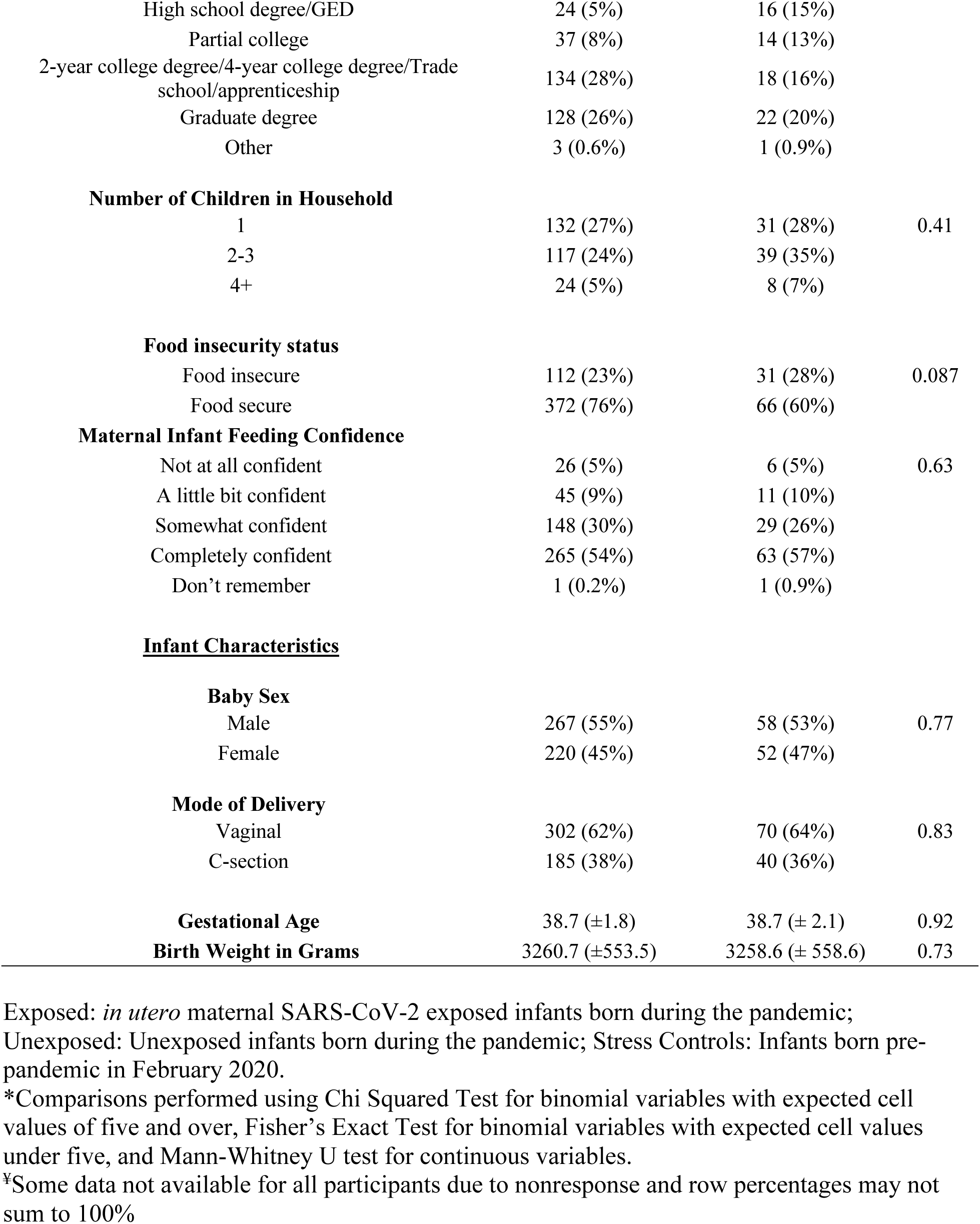
Demographic Characteristics between COMBO Study Waves.

